# Factors Associated With Childhood Immunization Coverage among Children Aged 0–23 Months in the Oyibi Sub-Municipality, Kpone-Katamanso Municipality, Ghana

**DOI:** 10.64898/2026.01.20.26344493

**Authors:** Michael Addipah

## Abstract

**Background:** Childhood immunization remains one of the most cost-effective public health strategies for reducing vaccine-preventable diseases (VPDs) and achieving Sustainable Development Goal 3. Despite improvements in global and national immunization coverage, significant gaps persist in Ghana, particularly in peri-urban districts such as Kpone-Katamanso. This study assessed factors associated with childhood immunization coverage among children aged 0–23 months in the Oyibi sub-municipality.

**Methods:** A community-based cross-sectional study was conducted among 360 mothers/caregivers attending Child Welfare Clinics (CWCs) across six communities in Oyibi. A structured questionnaire was administered, and data were analyzed using STATA 15. Descriptive statistics were conducted, followed by chi-square and Fisher’s exact tests to assess associations. Logistic regression models were used to determine predictors of immunization coverage at a 95% confidence level.

**Results:** Overall, 66.7% of children received all required vaccines. However, 50.2% of mothers/caregivers had previously missed an immunization appointment. Mothers who ensured complete vaccination for their children had significantly reduced odds of missing immunization schedules (OR = 0.34; 95% CI: 0.21–0.56; p < 0.001). After adjusting for confounders, receiving all required vaccines was strongly protective against missing scheduled visits (AOR = 0.20; 95% CI: 0.09–0.44; p < 0.001). Immunization coverage was estimated at 50.4%, falling short of Ghana’s 95% target.

**Conclusion:** Immunization coverage in Oyibi remains suboptimal, with substantial defaulting on scheduled vaccination sessions. Strengthened community education, improved health-worker engagement, reduced waiting time, and timely vaccine availability are critical for improving immunization uptake.

## INTRODUCTION

Childhood immunization is recognized globally as one of the most effective public health interventions for reducing morbidity and mortality from vaccine-preventable diseases (VPDs). Since the introduction of the Expanded Programme on Immunization (EPI) in 1974, global immunization has dramatically improved, contributing to reductions in childhood deaths and advancing efforts toward achieving the Sustainable Development Goals (SDGs) [1]. Immunization directly supports SDG 3, which aims to reduce under-five mortality to fewer than 25 deaths per 1,000 live births by 2030 [2].

Despite global gains, disparities in immunization coverage remain pronounced, particularly in low- and middle-income countries (LMICs). In Africa, DPT1 and DPT3 coverage (84% and 76%, respectively) still lag behind coverage in more developed regions [3]. Ghana has made notable progress, with childhood vaccination rates increasing from 47% in 1988 to 73% in 2022 [4]. However, national-level improvements mask regional and district-level disparities, especially in peri-urban and fast-growing municipalities such as Kpone-Katamanso.

Oyibi sub-municipality, characterized by rapid urbanization and high population mobility, experiences challenges with achieving consistent immunization uptake. The municipal health report indicates that only 60.3% of eligible children were fully immunized in 2022, far below Ghana’s target of 95% full immunization coverage [5]. Factors such as vaccine hesitancy, caregiver socio-demographics, socio-cultural beliefs, health system challenges, and accessibility barriers contribute to these gaps [6–9].

Vaccine hesitancy—defined as a delay in acceptance or refusal despite availability of vaccination services—continues to undermine immunization efforts globally [10]. In Ghana, caregiver beliefs, religious influences, misconceptions about vaccine safety, and negative interactions with health workers have been documented as important contributors to missed vaccination opportunities [11–13]. Additional structural challenges, such as long waiting times, staff shortages, and inconsistent vaccine availability, further impede uptake [14].

Given these persistent challenges and the need for localized evidence to support municipal-level immunization strengthening, this study assessed factors associated with childhood immunization coverage among children aged 0–23 months in the Oyibi sub-municipality.

## METHODS

### Study Design

A community-based cross-sectional design was used to assess mothers’ and caregivers’ factors associated with immunization coverage within the six communities in the Oyibi sub-municipality.

### Study Area

The study was conducted in the Oyibi sub-municipality of Kpone-Katamanso, Greater Accra Region of Ghana.

### Population

Mothers/caregivers with children aged 0–23 months attending CWCs within the six communities in the municipality.

### Sample Size and Sampling

A sample size of 360 was obtained using the Krejcie & Morgan formula. Probability sampling with systematic random selection was used across the six communities.

### Data Collection

Structured questionnaires captured information on socio-demographic characteristics, accessibility, socio-cultural perceptions, and health facility experience.

### Data Analysis

Data were analyzed in STATA 15.0. Descriptive statistics were summarized using frequencies. Associations were assessed using chi-square or Fisher’s exact test. Logistic regression produced crude and adjusted odds ratios with 95% CI.

### Ethical Considerations

Ethical approval was obtained from the Ghana Health Service Ethics Review Committee.

### Funding

This study did not receive any funding from any organization, institution and agencies in the public, private nor commercial space.

## RESULTS

A total of 360 mothers/caregivers participated in the study. Most respondents (50.0%) were aged 26–35 years, followed by 36–45 years (29.2%). The majority were married (72.2%), Christians (90.8%), and employed (55.8%). More than half (60.0%) had attained at least secondary education, and 35.3% reported monthly income between GH¢500–900.00

### Association Between Socio-demographic Factors and Immunization Coverage

Bivariate analysis showed that certain socio-demographic variables were significantly associated with full immunization. Children whose mothers had secondary education were more likely to be fully immunized compared to those of primary-educated mothers (OR = 0.37; p = 0.059). Higher monthly income (GH¢2000–2400) had a strong positive association with immunization (OR = 0.25; p = 0.016).

### Association Between Child Characteristics and Immunization Coverage

Child age was significantly associated with immunization status. Children younger than seven months had the highest proportion of full immunization (65.79%). In contrast, children aged 13– 18 months had the lowest full immunization rate (36.84%), indicating missed opportunities as children grow older. These findings are shown in **Table 3**.

### Accessibility of Health Services and Immunization Status

Accessibility-related variables showed significant associations with immunization. Mothers who perceived road conditions as “somewhat good” were twice as likely to default compared to those with good roads. Poor transportation and distance barriers also contributed to reduced immunization uptake. See **Table 4** below.

### Reasons for Non-Compliance With the Immunization Schedule

The leading reason for non-compliance with the immunization schedule was fear of adverse drug effects, reported by 79% of caregivers. Other major barriers included financial constraints (58%), long-distance travel to facilities (54%), being too busy due to social engagements (47%), and long waiting times (44%). Concerns about vaccine safety accounted for 10%, while 4% cited other reasons. These findings highlight a combination of perceived side effects, economic challenges, accessibility issues, and service delivery bottlenecks as key drivers of missed immunization appointments.

### Health Service Provision Factors

Health system factors demonstrated notable influence on immunization uptake. Waiting time <30 minutes was significantly associated with full immunization (p = 0.010). Staff adequacy and staff attitude showed trends but were not statistically significant. **Table 5** provides details.

### Reasons for Waiting Time

The most frequently reported cause of prolonged waiting time was limited staff strength, cited by 26% (n = 94) of respondents. Other notable factors included high client turnout (16%; n = 58), nurses’ behavior during work (15%; n = 54), and instances where caregivers experienced no delay (15%; n = 53). Additional contributors were vaccine delays (10%; n = 36), lateness of nurses to work (9%; n = 33), and mothers’ own delays (8%; n = 30). A very small proportion (1%; n = 2) reported indifference or other unspecified reasons.

These findings highlight staffing shortages and high service demand as the primary drivers of long waiting times in immunization services.

### Socio-cultural Factors and Immunization Coverage

Sociocultural beliefs and spouse influence significantly affected immunization. Mothers whose spouses discouraged immunization had the lowest full-immunization rates (28.97%; p = 0.001). Negative religious or cultural perceptions were also linked with noncompliance. These findings are summarized in **Table 6**.

### Logistic Regression Results

Multivariate logistic regression identified **three independent predictors** of full immunization status:

1. Child age (7–12 months more likely not fully immunized)
2. Socio-demographic income level
3. Receipt of all required vaccines (strongest predictor)

Receiving all required vaccines reduced the odds of missing immunization by **80% (AOR = 0.20; p < 0.001)**.

Interpretation: Mothers whose children received all required vaccines had significantly reduced odds of missing scheduled immunization (AOR = 0.20).

## DISCUSSION

This study evaluated socio-demographic, cultural, accessibility, and health system determinants of immunization coverage among children aged 0–23 months in the Oyibi sub-municipality. Despite improvements in Ghana’s national immunization performance, this study found that only 50.4% of children met the criteria for complete immunization, a proportion lower than the national average and far from the 95% national target.

Consistent with previous studies, caregiver education, marital status, employment, and household income showed significant associations with immunization status. This aligns with findings from Ethiopia [15], India [16], and Senegal [17], where maternal education and socio-economic status were strong predictors of immunization uptake.

Health system factors emerged prominently. Long waiting times, irregular staff presence, and negative provider attitudes contributed to missed appointments. These findings corroborate studies in Ghana and Nigeria indicating that quality of interaction with health workers significantly influences caregiver adherence to immunization schedules [18, 19].

Socio-cultural determinants—particularly religious influence, myths surrounding vaccine safety, and spouse approval—also shaped immunization behaviors. These observations reflect global patterns outlined in the WHO Vaccine Hesitancy Matrix [10].

Overall, the findings highlight the need for multifaceted strategies, including community sensitization, improved service delivery, and targeted engagement of caregivers with low education or income levels.

## CONCLUSION

Immunization coverage in the Oyibi sub-municipality remains inadequate, with substantial proportions of caregivers missing scheduled appointments. Socio-demographic characteristics, accessibility barriers, health system constraints, and cultural beliefs all influence immunization behavior. Strengthened health education, improved clinic efficiency, and culturally sensitive community engagement are crucial to increasing immunization uptake.

## Data Availability

The dataset used for the study is available from the author on request. Permission will be requested from the Municipal Health Directorate of Kpone Katamanso and Ghana Health Service Ethics Review Committee before giving it out on request.

## Authors Details and Author contributions

Michael Addipah

Volunteer for Expanded Program on Immunization EPI Ghana Since 2012

Kpone Katamanso Municipal Health Service, Ghana Health Service

City: Regional Capital, Accra Ghana Country: Ghana

Degree: Master of Public Health with emphasis on Epidemiology and Disease Control

School: University of Ghana, Legon College of Health Sciences, School of Public Health

Conceptualization: Michael Addipah.

Data curation: Michael Addipah.

Formal analysis: Michael Addipah.

Investigation: Michael Addipah

Methodology: Michael Addipah.

Project administration: Michael Addipah

Resources: Michael Addipah

Software: Michael Addipah.

Supervision: Michael Addipah.

Validation: Michael Addipah.

Visualization: Michael Addipah.

Writing – original draft: Michael Addipah.

Writing – review & editing: Michael Addipah

**Fig. 1.**
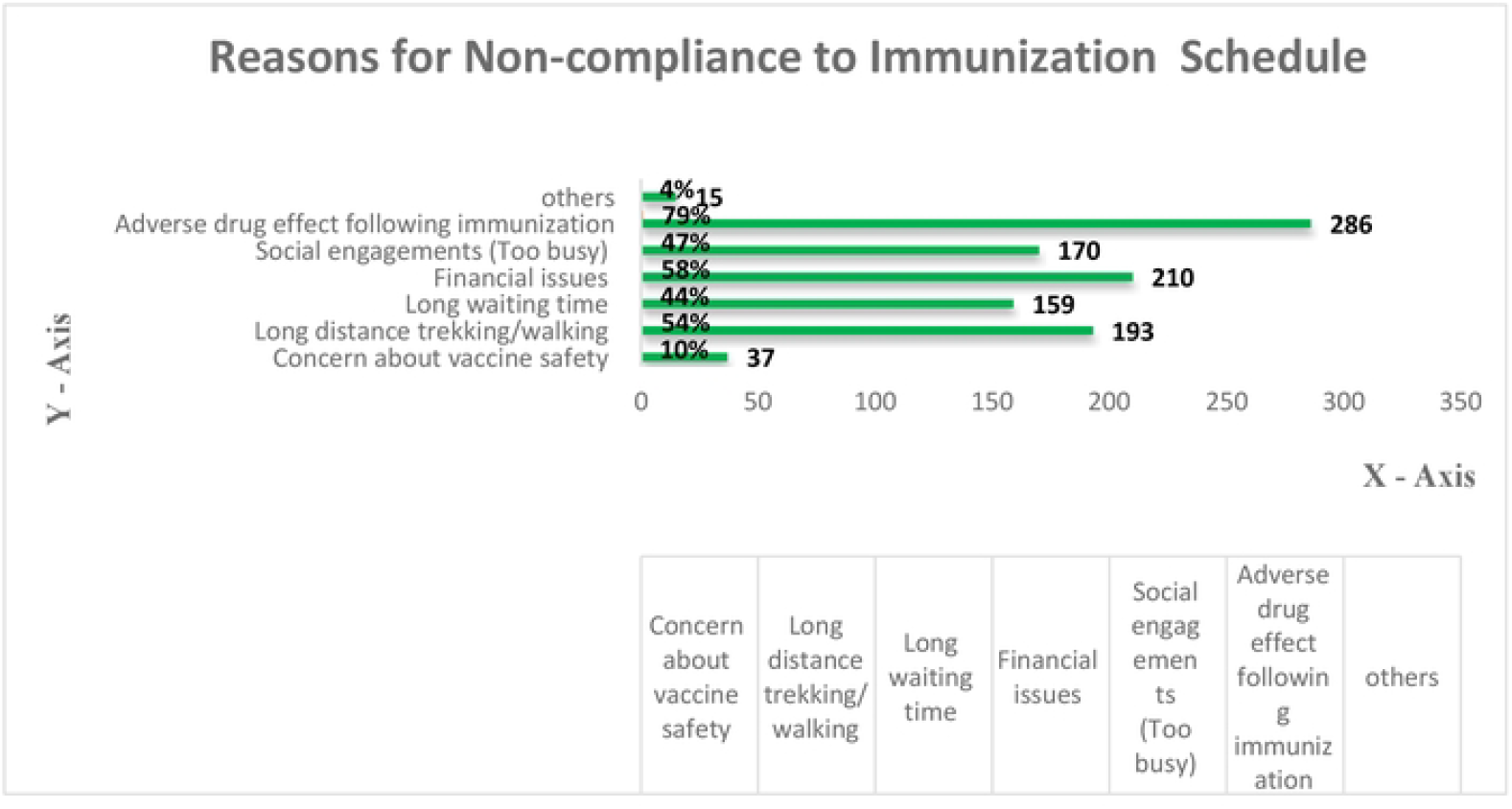

**Fig. 2.**
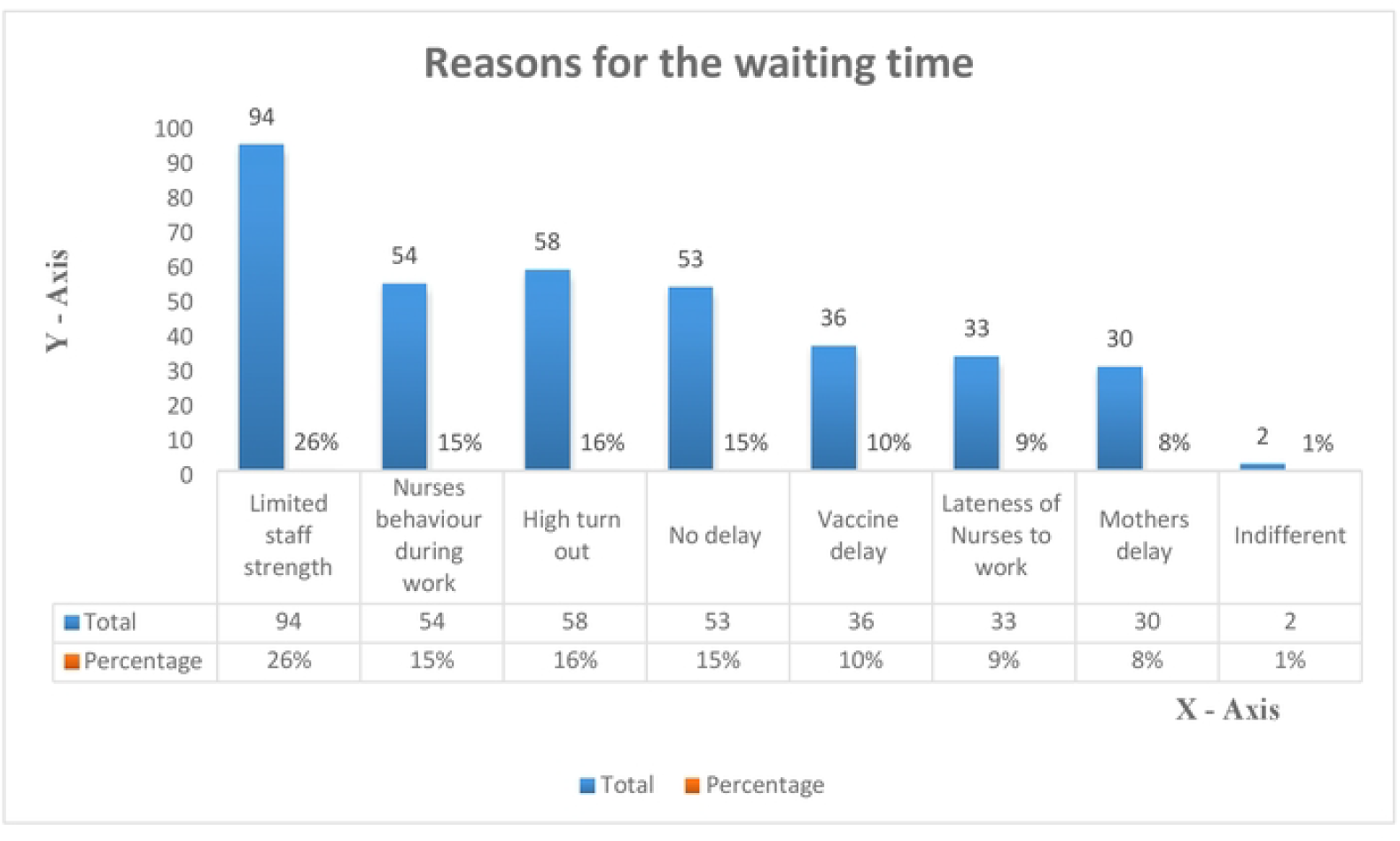

## Notes

### Competing Interest Statement

The authors have declared no competing interest.

### Funding Statement

The survey did not receive any financial support from anybody. it was soley the funded by the Researcher who is also the principal investigator.

### Author Declarations

Ghana Health Service Ethics Review Committee, Ghana. Kpone Katamanso Municipal Health Directorate, Ghana

## References

1. Abdulraheem LS, Onajole AT, Jimoh AAG, Oladipo AR (2011) Reasons for incomplete vaccination and factors for missed opportunities among rural Nigerian children. J Public Health Epidemiol 3(4): 194–203.

2. Adedokun ST, Uthman OA, Adekanmbi VT, Wiysonge CS (2017) Incomplete childhood immunization in Nigeria: A multilevel analysis of individual and contextual factors. BMC Public Health 17: 236. 10.1186/s12889-017-4137-7

3. Al-Moukhtar O, Al O (2011) Knowledge, attitude and practices of mothers regarding immunization of infants and preschool children at Al-Beida City, Libya 2008. Egypt J Pediatr Allergy Immunol 9(1): 29–34.

4. Armah G, Fuentes S, Korpela KE, Harris VC, Parashar U, Victor JC, et al. (2016) Significant correlation between the infant gut microbiome and rotavirus vaccine response in rural Ghana. J Infect Dis 215(1): 34–41.

5. Asuman D, Ackah CG, Enemark U (2018) Inequalities in child immunization coverage in Ghana: Evidence from a decomposition analysis. Health Econ Rev 8(1): 9–12.

6. Babalola S (2011) Maternal reasons for non-immunization and partial immunization in northern Nigeria. J Paediatr Child Health 47: 276. 10.1111/j.1440-1754.2010.01956.x

7. Brearley L, Eggers R, Steinglass R, Vandelaer J (2013) Applying an equity lens in the Decade of Vaccines. Vaccine 31(Suppl 2): B103–B107. 10.1016/j.vaccine.2012.11.088

8. Centers for Disease Control and Prevention (CDC) (2006) Vaccine-preventable deaths and the Global Immunization Vision and Strategy, 2006–2015. MMWR Morb Mortal Wkly Rep 55(18): 511–515.

9. Tauil MC, Sato AP, Waldman EA (2016) Factors associated with incomplete or delayed vaccination across countries: A systematic review. Vaccine 34(24): 2635–2643. 10.1016/j.vaccine.2016.04.016

10. UNICEF (2015) The State of the World’s Children 2014: Child survival. New York: United Nations Children’s Fund.

11. World Health Organization (2022) Immunization agenda 2030: A global strategy to leave no one behind. Geneva: WHO. Available from: https://cdn.who.int/media/docs/defaultsource/immunization/strategy/ia2030/

12. World Health Organization (2021) World health statistics. Geneva: WHO.

13. World Health Organization (2018) World health statistics 2018: Monitoring health for the SDGs. Geneva: WHO.

14. World Health Organization (2005) Global Immunization Vision and Strategy 2004–2015. Geneva: WHO. Available from: http://www.who.int/vaccinesdocuments/DocsPDF05/GIVSFinalEN.pdf

15. World Health Organization (2005) WHO fact sheet no. 288. Geneva: WHO. Available from: http://www.who.int/mediacentre/factsheets/

16. World Health Organization Regional Office for Europe (2014) European Vaccine Action Plan 2015–2020. Copenhagen: WHO Regional Office for Europe.

17. Williams SE (2014) What are the factors that contribute to parental vaccine hesitancy and what can we do about it? Hum Vaccin Immunother 11(5): 551–556. 10.4161/hv.28596

18. Ministry of Health (2014) Immunization programme comprehensive multi-year plan in line with Global Immunization Vision and Strategies. Accra: Ministry of Health, Ghana.

19. Perez F, Ba H, Dastagire SG, Altmann M (2009) The role of community health workers in improving child health programmes in Mali. BMC Int Health Hum Rights 9(1): 28. 10.1186/1472-698X-9-28

